# Modeling on Wastewater Treatment Process in Saudi Arabia: a perspective of Covid-19

**DOI:** 10.1101/2021.11.22.21266599

**Authors:** Abdullah Ahmadini, Ahmed Msmali, Zico Mutum, Yashpal Singh Raghav

## Abstract

The novel coronavirus disease (COVID-19) pandemic has had devastating effects on healthcare systems and the global economy. Moreover, coronavirus has been found in human feces, sewage, and in wastewater treatment plants. In this paper, we highlight the transmission behavior, occurrence, and persistence of the virus in sewage and wastewater treatment plants. Our approach follows the process of identifying a coronavirus hotspot through existing wastewater plants in major cities of Saudi Arabia. The mathematical distributions, including the log-normal distribution, Gaussian model, and susceptible exposed infected recovery (SEIR) model, are adopted to predict the coronavirus load in wastewater plants. We highlight not only the potential virus removal techniques from wastewater treatment plants, but also methods of tracing SARS-CoV-2 in humans through wastewater treatment plants.

## 1. Introduction

The contamination of wastewater with viruses causing various diseases is an emerging threat that is increasing owing to the effects of climate change [35]. With the outbreak of the COVID-19 pandemic, contamination of water with viruses has become an issue of concern. The coronavirus can be transmitted from wastewater to workers in wastewater treatment plants. However, a hot climate may reduce the persistence of the virus in sewage systems; thus, the spread of the virus in such climates may not be significant [39].

Globally, researchers are testing wastewater for the novel coronavirus to estimate the total number of infections in a community, given that most people will not be tested. Coronavirus can be detected in feces within three days of infection, which is significantly earlier than the time when people begin to show severe symptoms and seek medical care [33]. Therefore, wastewater testing helps assess the spread of coronavirus [36]. Coronavirus has been correlated with viral RNA, which can be detected using real-time quantitative reverse transcription polymerase chain reaction PCR (RT-qPCR). The occurrence of the virus in wastewater has been found to positively correlate (correlation coefficient *r* > 0.9) with local hospital admissions [34].

On March 2, 2020, the Ministry of Health in Saudi Arabia announced the first case of COVID-19. By April 9, 2020, the number of COVID-19 confirmed cases in the country had increased to 2932. This number increased rapidly in June and July 2020. Recently, however, the number of new confirmed cases is gradually reducing. From January 3, 2020, to November 12, 2021, there were 548,973 COVID-19 confirmed cases in Saudi Arabia, resulting in 8,805 deaths. A total of 46,288,357 persons had been vaccinated within that period [51]. The wastewater approach for estimating the spread of coronavirus has scarcely been used in Saudi Arabia. A few studies [37–39] have been conducted on wastewater surveillance to track COVID-19 in Saudi Arabia. In this study, we investigate some models that will aid our understanding of the occurrence and persistence of coronavirus in wastewaters. Using a mathematical distribution, including normal distribution, log-normal distribution, and the susceptible exposed infected recovery (SEIR) epidemiology model, we suggest a possible model that can be used to identify coronavirus hotspots by analyzing wastewaters.

In this paper, Section 2 highlights wastewater treatment plants in Saudi Arabia and their models. Section 3 focuses on the techniques used for viral removal from wastewater treatment plants. In Section 4, we provide the material and methods used for tracing the coronavirus in sewage and untreated wastewater plants, and Section 5 provides the discussion and results of our work. Finally, Section 6 presents the conclusions of our study.

## 2. Wastewater and Models

Saudi Arabia has witnessed one of the fastest socio-economic developments over the last few decades with increased urbanization. Consequently, water demand for all purposes has tremendously increased in a country with a hot climate and limited natural water resources. According to *Qatrah* [4], Saudi Arabia is the world’s third-largest per capita consumer of water after the United States. This gap between water demand and supply is filled by groundwater resources, water desalination, and the economical use of treated wastewater. In Saudi Arabia, wastewater treatment plants are, therefore, essential to bridge this gap. Treated wastewater is considered an important part of water resources in Saudi Arabia and is used as a major source for non-potable water demands, such as agricultural, industrial, and commercial uses [5]. The Ministry of Water and Electricity aims to provide entire sewage collection and treated wastewater services to every city with a population above five thousand by 2025 [6].

Currently, there are thirty-three wastewater treatment plants in Saudi Arabia with a total capacity of 748 million cubic meters per year. Fifteen more are under construction. Water reuse in Saudi Arabia is growing in both urban and rural areas. Treated wastewater is mainly used for landscaping, irrigation, and in industries such as refining. In Riyadh, the capital city of Saudi Arabia, 50 million cubic meters of treated wastewater is pumped over 40 km and elevated over 60 km to irrigate about 15,000 hectares of land per year [7]. The major wastewater treatment plants in Saudi Arabia serving various cities are listed in Table 1. The table shows the insufficiency of the wastewater management system in Saudi Arabia.

**TABLE 1:** Major wastewater treatment plants in Saudi Arabia (source: King Abdullah University of Science and Technology).

| Wastewater treatment plants: number and capacity (1000 m<sup>3</sup>/day) |  |  |  |  |  |  |  |  |  |  |
| --- | --- | --- | --- | --- | --- | --- | --- | --- | --- | --- |
| Region | Calculated wastewater flow capacity (1000 m<sup>3</sup>/day) |  |  | Existing |  | Under construction |  | Planned |  | Future capacity (by year 2035) |
|  | 2010 | 2025 | 2035 | No. | Cap. | No. | Cap. | No. | Cap. |  |
| Al Baha | 56 | 81 | 107 | 0 | 0 | 1 | 16.2 | 6 | 72.6 | 107 |
| Al Jouf | 69 | 96 | 119 | 2 | 38 | 2 | 57.5 | 1 | 22 | 119 |
| Assir | 294 | 419 | 529 | 4 | 82.5 | 7 | 172.5 | 0 | - | 529 |
| Eastern Province | 648 | 905 | 1128 | 13 | 527.3 | 6 | 251 | 1 | 400 | 1128 |
| Hail | 93 | 130 | 162 | 1 | 19.2 | 2 | 87.6 | 1 | 55.8 | 162 |
| Jizan | 203 | 301 | 381 | 1 | 20 | 0 | - | 22 | 112 | 381 |
| Madinah | 275 | 390 | 496 | 4 | 351 | 5 | 34 | 0 | - | 496 |
| Makkah | 1100 | 1532 | 1911 | 15 | 888 | 6 | 902 | 1 | 113 | 1911 |
| Najran | 78 | 107 | 133 | 0 | 0 | 1 | 60 | 5 | 170 | 133 |
| Northern Borders | 47 | 71 | 88 | 2 | 24 | 1 | 24 | 1 | 25 | 88 |
| Qaseem | 185 | 258 | 328 | 5 | 131.5 | 3 | 125 | 0 | - | 328 |
| Riyadh | 1066 | 1513 | 1890 | 10 | 993.5 | 7 | 443.5 | 2 | 530 | 1890 |
| Tabouk | 120 | 169 | 214 | 1 | 60 | 1 | 15 | 0 | - | 214 |
| Total | 4234 | 5972 | 7486 | 3135 |  | 2188.3 |  | 1500.4 |  | 7486 |

The most commonly used secondary treatment mechanism for wastewater in Saudi Arabia is the conventional activated sludge system, as shown in Table 2. Other common treatment techniques are filtration and chlorination; in addition, a few wastewater treatment plants use reverse osmosis. A major challenge of wastewater treatment plants (WWTPs) in Saudi Arabia is the lack of proper connectivity between the sewage system and existing plants.

**TABLE 2:** Waste water treatment plants, working mechanism and capacity in Saudi Arabia.

| WWTPs | Treatment Process | Capacity ($\frac{m^3}{year}$) | Treatment Level | Purpose |
| --- | --- | --- | --- | --- |
| Manfouha-Riyadh | Trickling filter, activated sludge | North 200,000, South 200,000, East 200,000 | Tertiary | Irrigation |
| Heet-Alkharj | Activated sludge | Phase I 100,000, Phase II 100,000, Phase III (under construction) 200,000 | Tertiary | Irrigation, groundwater recharge |
| Hayer-Riyadh | Activated sludge | Phase I 400,000 | Tertiary | Irrigation, groundwater recharge |
| Refinery-Riyadh | Clarification and filtration | 20,000 | Tertiary | Agricultural irrigation |
| Dammam | Activated sludge | 215,000 | Tertiary | Landscape irrigation |
| Al Madinah | Media filtration | 460,000 | Tertiary | Agricultural irrigation |
| Taif | Activated sludge | 190,000 | Tertiary/secondary | Landscape irrigation |
| Makkah | Trickling filter/activated sludge | Phase I 24,000, Phase II 50,000 | Tertiary | Irrigation and industrial |
| Qatif | Oxidation ditch | 210,000 | Tertiary | Landscape |
| Heet-Alkharj | Activated sludge | Phase I 100,000, Phase II 100,000, Phase III (under construction) 200,000 | Tertiary | Irrigation, groundwater recharge |
| Jizan Economic city | Activated sludge | 5400 | Tertiary | Irrigation and industrial |
| Alkhumrah, Jeddah | Activated sludge | 250000 | Tertiary | Landscape irrigation |

The knowledge and understanding of biological and chemical wastewater treatment include both advanced and conventional techniques. Advanced techniques include the application of mathematics, statistics, physics, chemistry, and bioprocess engineering. An overview of the sequence of wastewater treatment processes adopted by most of the existing wastewater treatment plants is shown in Figure 1.

**FIGURE 1:**
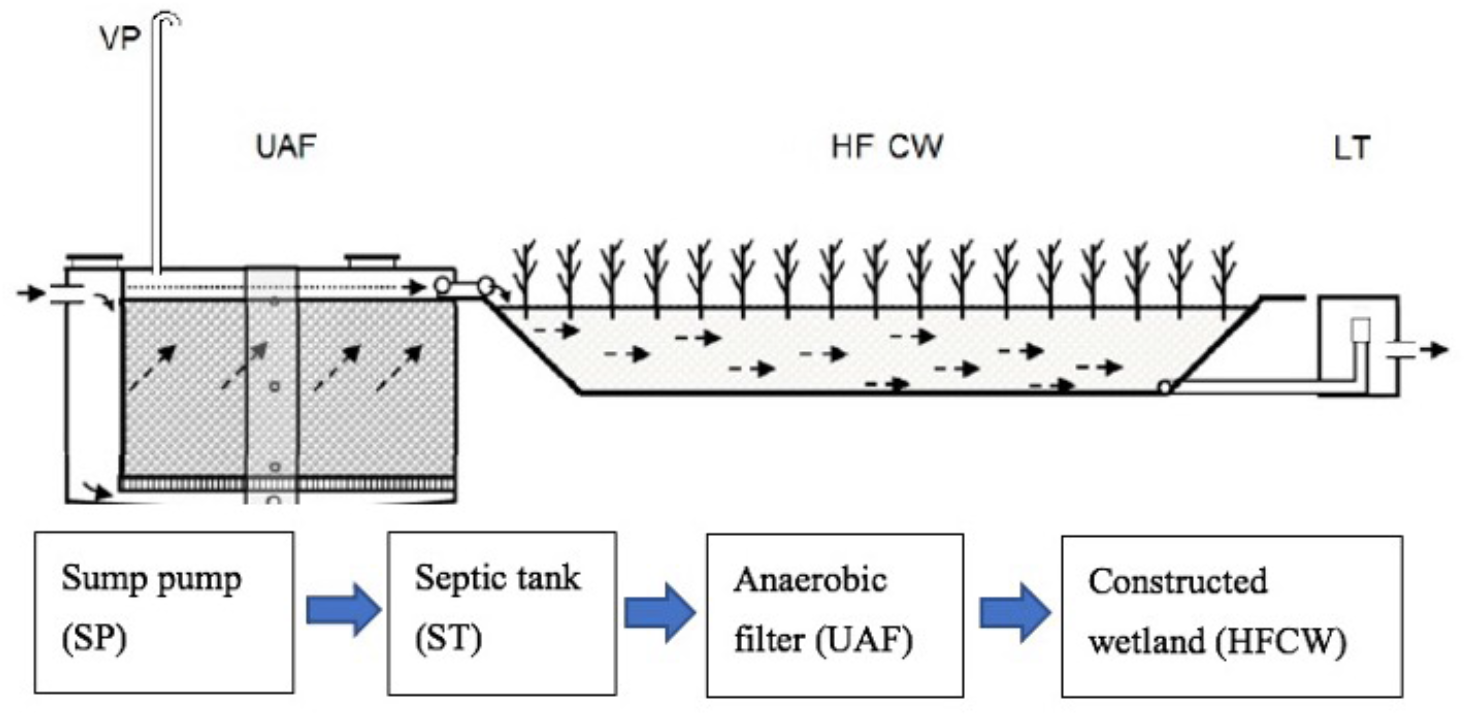
Important parts of the wastewater treatment process: vent pipe (VP), up-flow anaerobic filter (UAF), horizontal subsurface flow constructed wetland (HFCW), and level tank (LT) [9].

The wastewater treatment process consists mainly of a sump pump (SP), septic tank (ST), anaerobic filter (UAF), and constructed wetlands (HFCW). The purpose of the sump pump is to remove the coarse solids in the wastewater and pump the water into a two-chamber septic tank. Wastewater flows by gravity through an anaerobic filter and then through the constructed wetlands. Finally, the level tank (LT) controls the water level in the constructed wetland. Wetlands can improve wastewater quality by filtering before being released to open water.

The most widely used mathematical model for full-scale wastewater treatment plants is the activated sludge model No.1 (ASM1) [1]. The ASM1 model describes the process for organic and nitrogen removal from wastewater and validates its optimal performance using statistical models. An advanced mechanical description of the biological processes is provided in the updated version of the ASM model. The activated sludge model is one of the most important treatment processes for various wastewaters, and approximately 90% of municipal wastewater treatment plants use it in their treatment process [2,3]. Mathematical modeling of the wastewater treatment process plays an important role in the management of an effective treatment technique for existing plants.

In this section, we briefly illustrate some of the mathematical models used to study the quality of treated wastewater. First-order kinetic mathematical models are commonly used to compare contaminant removal efficiency and mass reduction in wastewater treatment plants. It is a nonlinear model used to predict pollutant removal in wastewater treatment systems [8]. The model equation is as follows.

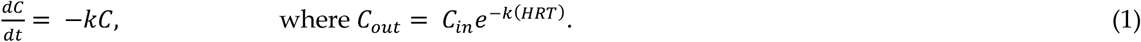

*C*_*out*_ is the concentration of the pollutant at the system outlet (mg/L), *C*_*in*_ is the concentration of the pollutant at the system inlet (mg/L), and HRT is the duration of hydraulic retention time of the system expressed in days. The monitoring data are adjusted to generate equation (1), and a similar process for this treatment is mentioned in [9]. The first-order kinetic model equation provides the parameter *k*, representing the rate of removal of pollutants or contaminants from wastewater.

The second-order kinetic mathematical model was used to predict the substrate removal from the wastewater treatment system as follows:

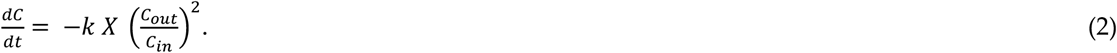

After linearization, we obtain

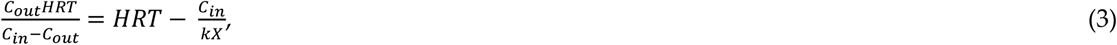

where 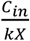 is the model constant.

Data adjustment can be performed for the wastewater treatment system for each step by nonlinear least squares. The statistical significance of each parameter model is measured by *p* value, and *R*^2^ is estimated to measure the goodness of fit to validate the model.

Multiple linear regression is another commonly used model to study the relationship between the dependent variable and independent variables and is also known as a predictor [10]. The different water quality parameters measured were used as the independent variables, and parameter for the water quality at the outlet were considered as the dependent variables. The equation for this model is given as *Y* = *αX*_1_ + *βX*_2_ + *γX*_3_, where Y is the independent variable and X is the dependent variable. Analyzing the multiple regression equations provides useful information for different water quality outflows.

## 3. COVID-19 and Wastewater

The occurrence of coronavirus in wastewater has shown the risk of persistent and fatal viral pandemics. Wastewater-based epidemiology (WBE) is a promising tool for assessing COVID-19 [15]. Currently, several applications of WBE exist, including the detection of illicit drugs, surveillance of pathogenic enteric viruses, and assessment of potential industrial chemical exposures [16]. Some viruses released from the feces of infected human are able to survive for days to months in aqueous environments [17]. For instance, SARS-CoV-2 can survive for more than 17 days at 4 °C and three days at 20 °C in hospital sewage, municipal sewage, and chlorine-free tap water [18]. Therefore, the occurrence of coronaviruses in wastewater is of significant concern. It has been reported that sewage contributes to SARS-CoV-2 transmission in urban regions. Later, it reinforced the potential for secondary transmission of the disease through wastewater, especially in regions that have unsatisfactory health and sanitation infrastructure [40].

Wastewater is one of the tools to help with surveillance of COVID-19 and its testing could be used as an early warning sign if the virus returns [30]. Several articles [31-33] have published confirming the detection of SARS-CoV-2 in sewage. It is necessary to implement proper disinfection of sewage to void infection with virus. Figure 2 illustrates the possible aspects of coronavirus contamination in treated wastewater.

**FIGURE 2.**
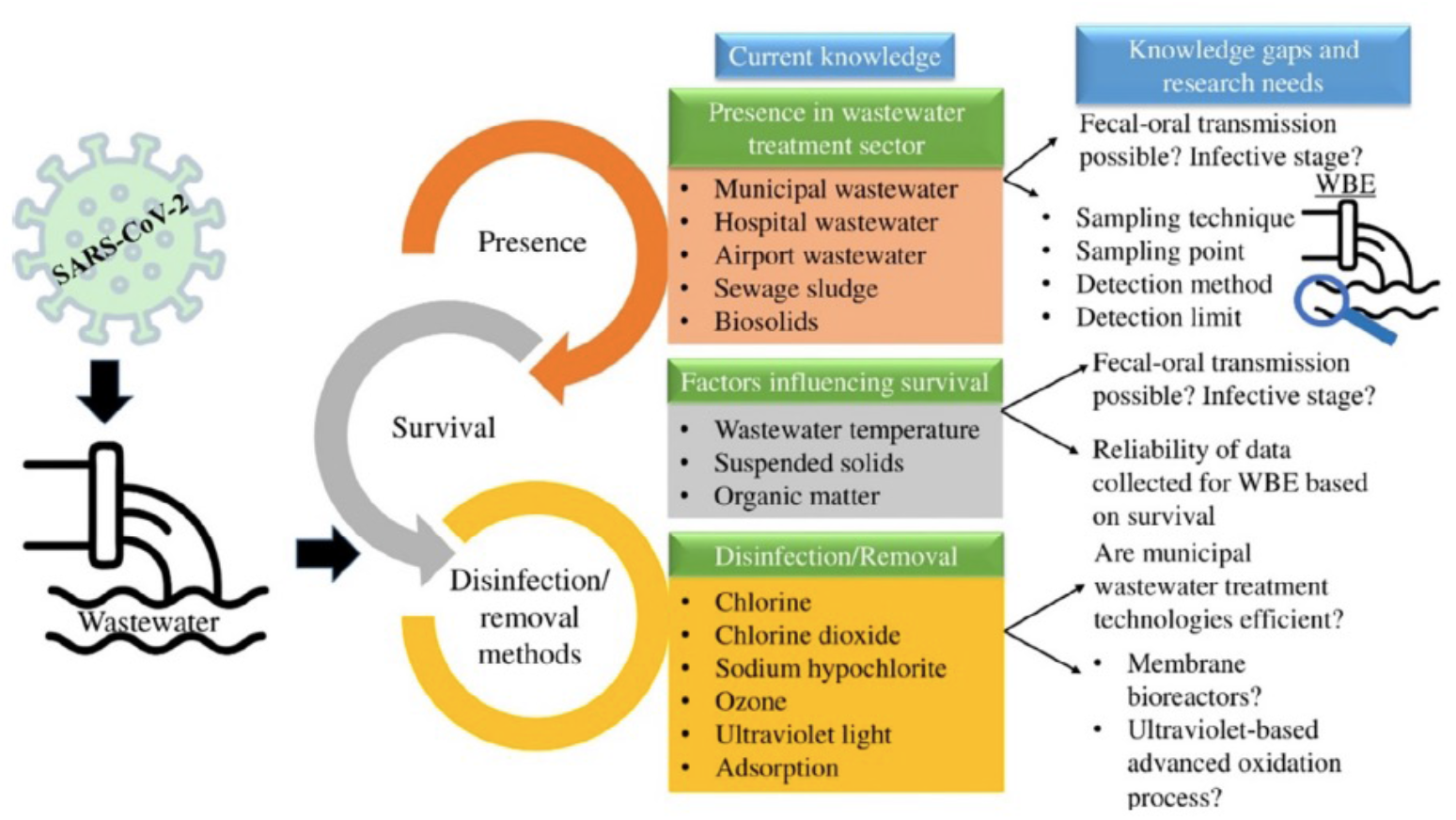
Features of SARS-CoV-2 in wastewater [39].

### 3.1 Ultraviolet (UV) and chlorine treatment

One of the most commonly used wastewater disinfection process includes the use of ultraviolet (UV) and chlorine. The disinfection process achieved 99.99% inactivation of fecal coliform; the fecal coliform concentration in the disinfected effluent was below detection (detection limit is 2 CFU/L) [19]. They observed that no COVID-19 were detected in the effluent after disinfection. The disinfection efficiency with chlorine is recommended by applying a dose higher than 6.5 mg/L and a contact time of at least 1.5 hours. It is more efficient to use UV radiation for disinfection of wastewater in hospitals that care for patients with COVID-19 due to the low amount by-products [20]. According to the WHO, the presence of residual chlorine of 0.5mg/L is standard and should be guaranteed in all water systems.

### 3.2 Ozone treatment

Ozone is widely used for the disinfection of many viruses in wastewater. According to International Ozone Association (IOA), research and testing has not been conducted on coronavirus. Therefore, a concrete conclusion cannot be made related to the disinfection of SARS-CoV-2 coronavirus by using Ozone [21]. Wastewater treatment using ozone could sufficiently disinfect viruses and provide a solid barrier to prevent them from entering the environment with the effluent, and ozone seems to be more effective than chlorine. There is a little information about the SARS-CoV-2 removal by using ozone disinfection processes, although the coronavirus is expected to be more sensitive to disinfection than other viruses. However, in [13], the authors reported that ozone could be one of the disinfectant methods for effectively inactivate SARS-CoV-2. Researchers at Tel Aviv University have demonstrated that ozone effectively sanitizes against coronavirus when exposed to low concentrations of gas [22].

The disinfection efficiency of the virus in wastewater depends on the ozone concentration and contact time. In [23], these values are calculated for specified log inactivation levels of a number of viruses in wastewater at pH 7.6 and 16 °C, with ranging within a specific value. Ozone is an efficient disinfection process as compared with conventional sewage treatment and able to reduce viral concentration by one to two log10 [11].

### 3.3 Sand filtration

Sand filtration is one of the most widely used wastewater treatment methods for wastewater treatment plants. One of the advanced wastewater treatment plants in Saudi Arabia is located in the north of Dhahran and uses 24 DynaSand filters for irrigation purposes [24]. Sand filtration is frequently used for the removal of dissolved particles and usually has less than 1 log 10 virus removal. However, in [56], the authors reported on the molecular mechanism for the unprecedented high virus removal using a practical sand filter. They developed functionalized sand filters using a water extract of Moringa oleifera seeds. They then tested the efficiency of the obtained sand using the MS2 bacteriophage virus, achieving a 7 log10 virus removal.

The viral removal activity of functionalized sand filter is due to protein binding. Some reported that the use of zero-valent iron sand filtration can remove virus contamination from wastewater. The zero-valent iron sand filters are efficiently able to remove some specific viruses such as MS2 and AiV(<1-2 log), but the removal efficiency varies among different viral species [54].

### 3.4 Membranes for virus removal

Water filtration membranes are categorized into four main types based on their pore size, they are: Reverse Osmosis (RO), Nanofiltration, Ultrafiltration and Microfiltration. Figure 3 shows the pore size and capabilities of these membranes for filtering particles, dissolved ions including salts, bacteria and pathogens.

**FIGURE 3.**
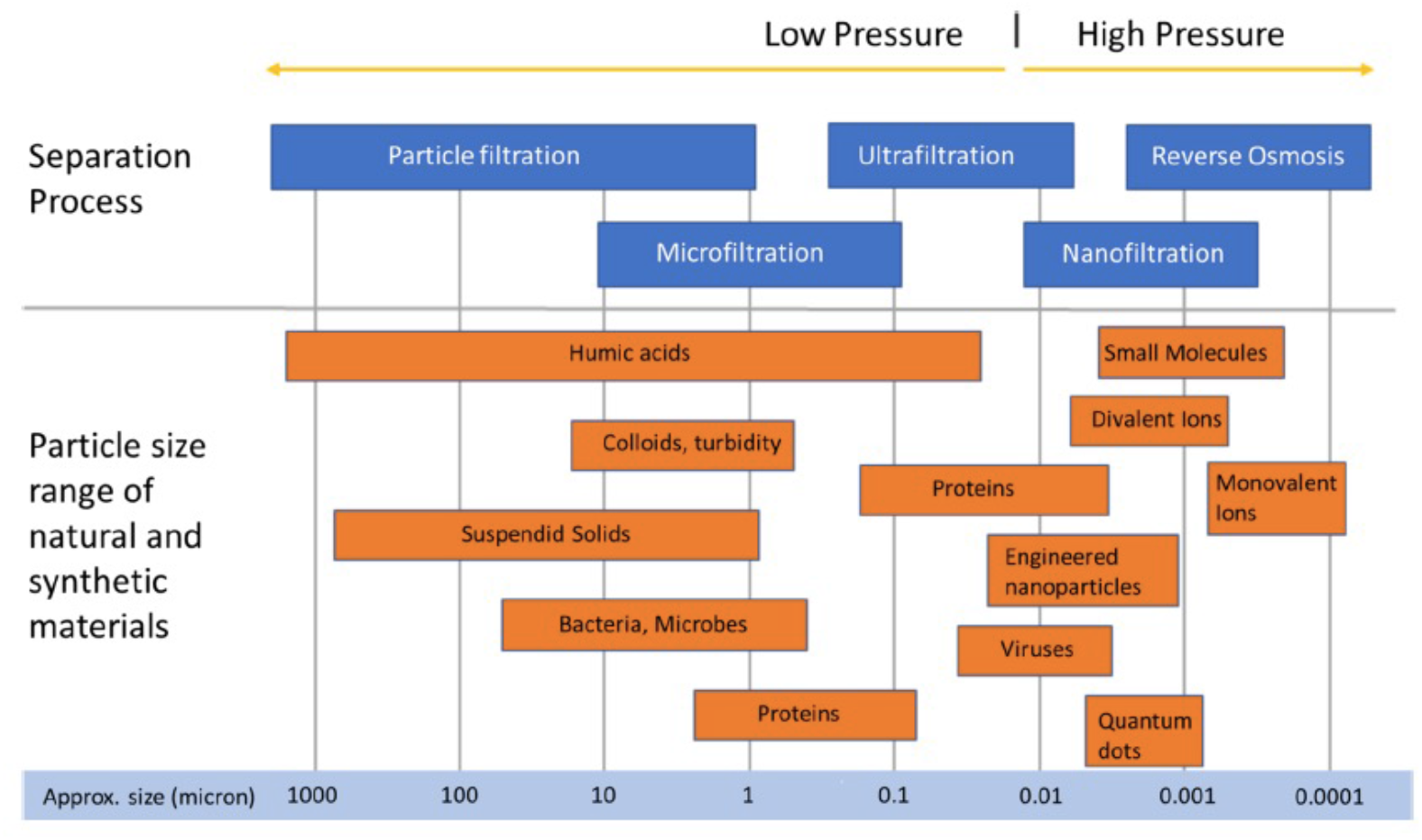
Comparison of water filtration membrane pore size with water contaminant size, reproduced from open source [12].

Since the diameter of SARS-CoV-2 is between 50 nm to 140 nm [47], membranes such as Reverse Osmosis, Nanofiltration and Ultrafiltration should be suitable for removing coronavirus from wastewater. In [57], the authors highlighted the use of polymeric and ceramic membranes for virus removal from water. It was reported that the viral removal efficiencies were highly variable since the range reported was 0.2–7.

The reverse osmosis and high-pressure nanofiltration membranes were popularly used for the treatment of water and desalination. The treatment process through these membrane systems was based on the solute transport by diffusion through the nonporous active layer of the membrane [14]. In this section, we outline some recent literatures on water filtration membranes used for the removal of viruses including SARS-CoV-2 from wastewater treatment plants.

#### 3.4.1 Reverse osmosis

Recently, in [41], the authors highlighted the treatment of waterborne pathogens by reverse osmosis. The reverse osmosis (RO) treatment method is considered as the last grade purification/treatment level. Any impurities with molecular weight greater than 200 are not able to pass through RO system. Similarly, the larger the ionic charge of the contaminants, the more likely to be rejected by RO membrane [60]. However, RO can be used in combination with other complementary processes such as pretreatment and posttreatment of particular matters to produce high quality recycled water [42]. In [61], the authors reported that the Membrane Bioreactor (MBR)/Reverse Osmosis (OR) systems were showing virus removal efficiencies (range of 2.3–2.9 log10). They stated that reverse osmosis or combined system (MBR/RO) were more advanced wastewater treatment systems with higher virus removal efficiency (>5 log10) for MS2 coliphages, whose virus size is smaller than SARS-CoV-2.

Szczuka et al. [43] reported that using forward-reverse osmosis for wastewater treatment had achieved higher than 98% rejection of organic contaminants, achieved at least 6.7 log10 removal of bacteriophage MS2 spiked into wastewater and achieved at least 5.4 log10 removal of native *E*.*Coil* in graywater and sewage. In [44], the authors used sand filters and reverse osmosis system for removing noroviruses of size 38-40 nm in diameter [45], from wastewater.

#### 3.4.2 Ultrafiltration

Ultrafiltration is often considered as one of the best pretreatment steps prior to reverse osmosis for the removal of human pathogens including bacteria, protozoa, and viruses from wastewater. Recently, in [46], the authors reviewed the use of ultrafiltration membranes for removing bacteria and viruses from wastewater. They reported that the use of ultrafiltration produced better water quality compared to conventional treatment due to the very fine pore structure. Using ultrafiltration for viral removal doesn’t require preconditioning of water samples, so it is widely used for water quality [49].

Ultrafiltration is a suitable membrane technique for removing coronaviruses with mean particle diameter of 120 nm and envelope diameter of 80 nm [48]. The adsorption property of solids in wastewater may facilitate the removal of coronavirus. It is also used for detection of viruses in wastewater. Three ultrafiltration devices: *the Centricon Plus -70, Amicon Ultra-15*, and *automatic Concentrating Pipette* have been successfully used to detect SARS-CoV-2 from wastewater [50].

#### 3.4.3 Nanomaterials

Nanotechnology and its application for water and wastewater treatment has emerged as a promising area and fast developing field. It has successfully used for the removal of viruses, bacteria, protozoans, and other contaminants from wastewater. The membranes with silver nanoparticles (AgNPs) have shown efficiency against MS2 bacteriophage virus and considered the best water treatment processes [52]. It is also reported that silver nanoparticles used in a nanocomposite filter system can be reached up to 100 percent removal of several different viruses. Electrospun nanofiber membranes (ENMs) are popularly used to treat pathogen contamination in wastewater. It is high flux and high rejection rate compared to conventional membranes, and offers a cost-effective, lightweight, and lower energy efficiency. ENMs are high porosity with approximately 80% while conventional membranes have 5-35% porosity [53]. Nanofibers containing tetraethoxysilane and ammonium tetrathiomolybdate mixed with polyacrylonitrile have enhanced more than 90% of viral removal efficiency. The use of nanoparticles alone or as a part of composite membranes could be more efficient in removing SARS-CoV-2 from wastewater [52].

#### 3.4.4 Microfiltration

Microfiltration membranes have average pore size and the viral removal from the wastewater treatment plant is lower compared to other membranes. In [58], the authors reported that the virus size (63 nm) was smaller than the microfiltration membrane size (0.2 μm), so it could pass through the membranes when the permeate flux was high or the membranes were clean. Moreover, in [59], they investigated that the pre-size of microfiltration was larger than 100 nm, which was more suitable for the removal of protozoa and bacteria rather than virus.

## 4. Material and Methods

We used the log-normal distribution, Gaussian model, and SEIR model to predict the infection of SARS-CoV-2 through wastewater. The theoretical assumptions and data considered in this study were collected and obtained from the Ministry of Health (MOH), Saudi Arabia website (https://covid19.moh.gov.sa/).

### 4.1 Normal Distribution

The normal distribution and its properties provide an important role in analytical investigation of the collection of coronavirus traces in untreated wastewater from wastewater treatment plants. Data of coronavirus traces collected from *N* wastewater treatment plants are assumed to follow a normal distribution. The rate of change in viral load at untreated wastewater plant with respect to time is given as follows;

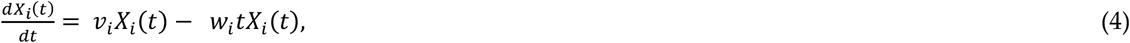

where *v*_*i*_ represents the change of mean and deviation due to daily new viral load; *w*_*i*_ represents the change of deviation of dataset related to the daily average viral load/concentration in untreated wastewater from *i* wastewater treatment plant. Let *X*_*i*_(*t*) be the viral load/concentration of coronavirus trance in untreated wastewater from *i* wastewater treatment plant, where *i* = 1,2,3,…, *N* during the time interval *t*_0_ ≤ *t* ≤ *T*. Let *μ* be the mean of viral load found in untreated wastewater from the wastewater treatment plant and *σ* be the related standard deviation. Then the above equation (4) can be written as follows;

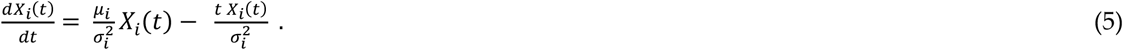

The exact solution of the above equation (5) is given as follows;

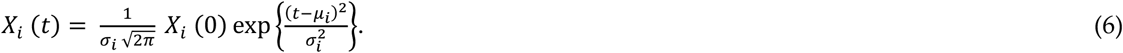

We can plot the set {*X*_*i*_(*t*): *i* = 1, 2, 3,……*N*} to forecast the possible viral load in untreated wastewater plants.

### 4.2 Log Normal Model

Log-normal distribution is an important statistical distribution to model many natural phenomena. A small percentage changes in the log-normal model correspond to numerous growth process and they are represented in log scales. If the effect of these changes is not significant, then the distribution will be closer to normal. We adopted the log-normal distribution to describe the spread of COVID-19 virus through wastewater treatment plants.

Suppose, the random change *R* is log-normal distribution, then the normal distribution *Z* = ln(*R*) follows the dynamics of the spread of COVID-19 virus. For instance, if *Z* = exp(*R*) has the behavior of the spread of the virus, then property of *R* = exp(*Z*) will be the log-normal distribution. In this case, the data collected from the wastewater treatment plants are assumed to follow log-normal distribution, then the mathematical equation is given as follows:

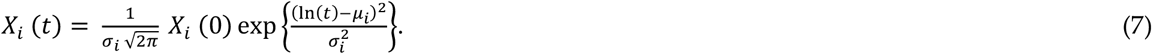

Comparing between the collected data and equation (7), we can determine the parameter *σ*_*i*_ as follows:

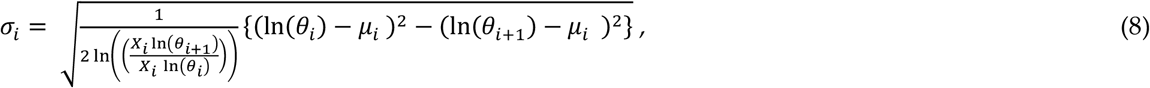

where {*θ*_*i*_: *i* = 1, 2, 3, 4,5} represents the number of days within the time interval *t*_0_ ≤ *t* ≤ *T*. We considered the collection of viral loads from untreated wastewater treatment plants for five consecutive days.

Figures 4 (a) and (b) show the normal equation (3) and the log-normal equation (5) for different viral loads. Considering that the collection from untreated wastewater plants was performed in five major cities (Riyadh, Makkah, Madinah, Jeddah, Dammam, Jazan), it is possible to observe high coronavirus traces owing to the high number of infected persons. Therefore, we chose different mean and standard deviation distributions, as shown in Figures 5 and 6. It is assumed that an adequate collection of data was performed. The data collected from the wastewater treatment plants in the major cities were plotted with respect to time. For instance, we assume that the data collected from each wastewater treatment plant from the respective major cities has a daily mean (*μ*_1_ = 0.2, *μ*_2_ = 0.3, *μ*_3_= 0.4, *μ*_4_ = 0.5, *μ*_5_= 2) and standard deviation (*σ*_1_ = 0.1, *σ*_2_ = 0.2, *σ*_3_ = 0.28, *σ*_4_ = 0.3, *σ*_5_ = 1) to be represented with the normal distribution.

**FIGURE 4.**
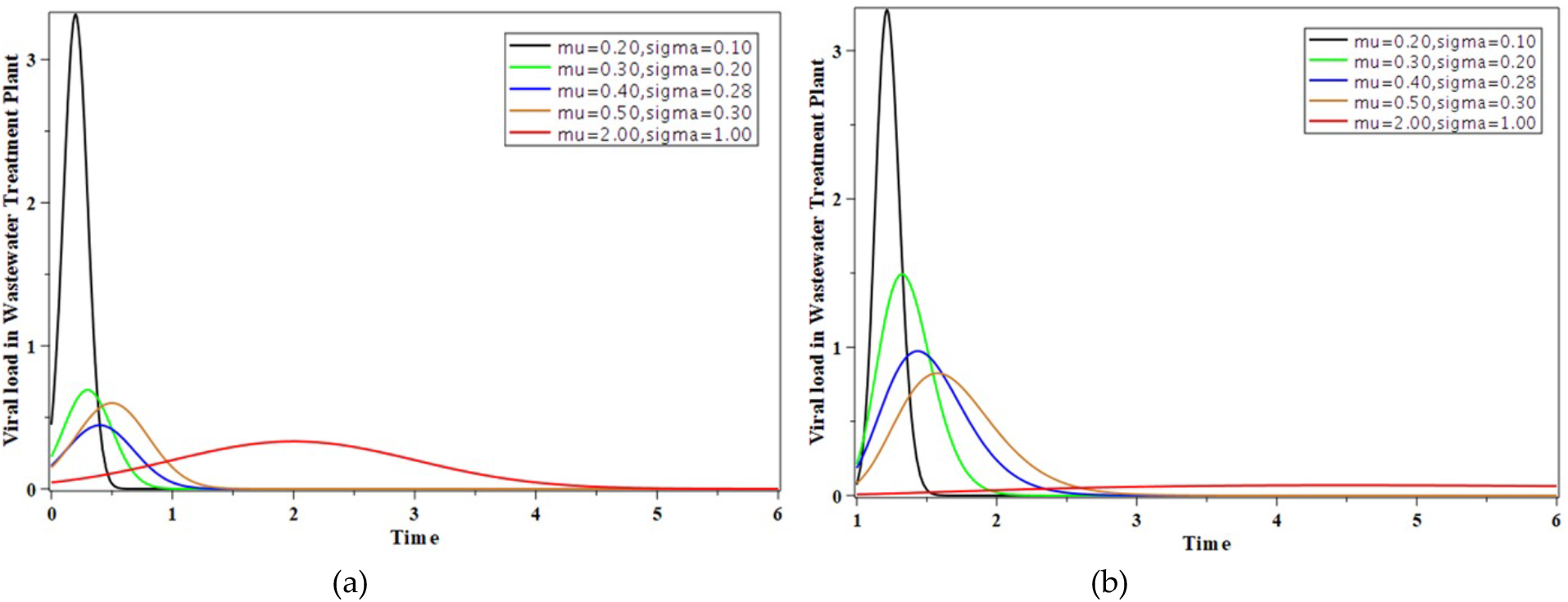
(a)Possible SARS-CoV-2 load in WWTP using Normal model, and (b) Possible SARS-CoV-2 load in WWTP using Log Normal model.

**FIGURE 5.**
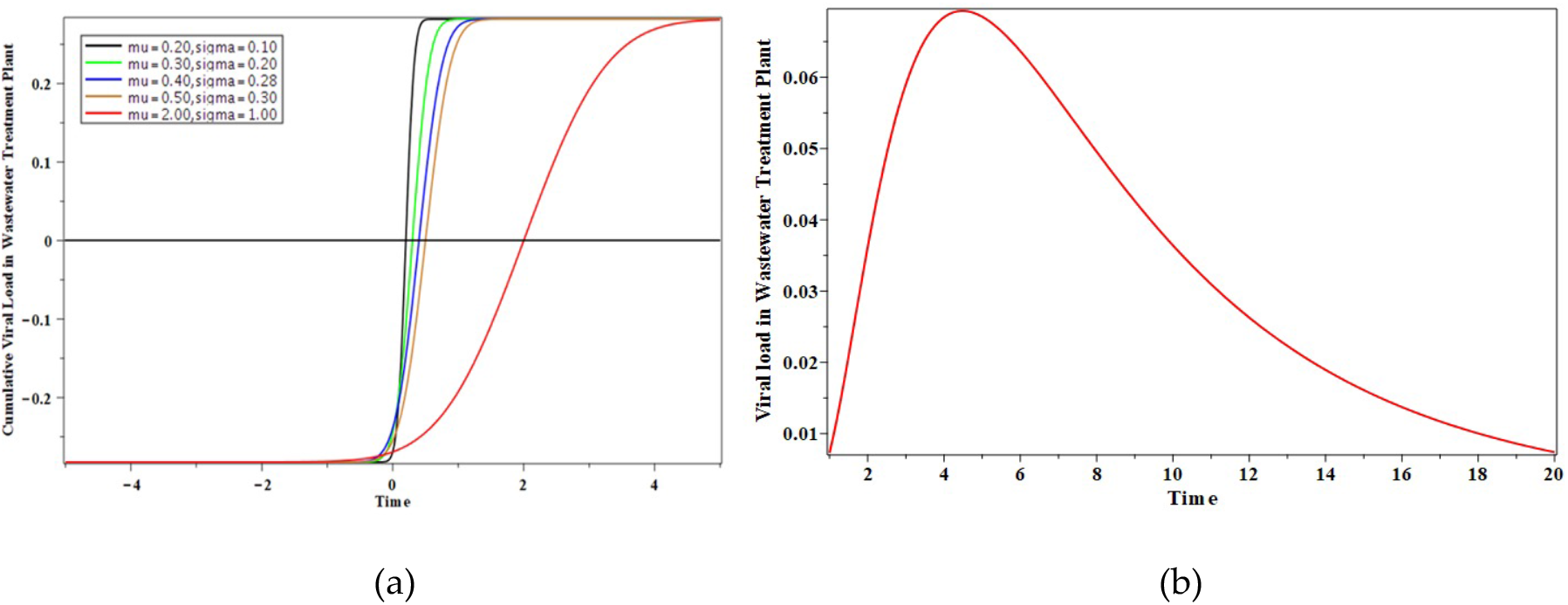
Gaussian model (a) predicting the possible cumulative SARS-CoV-2 load in WWTP, (b) predicting the possible SARS-CoV-2 load in WWTP.

**FIGURE 6.**
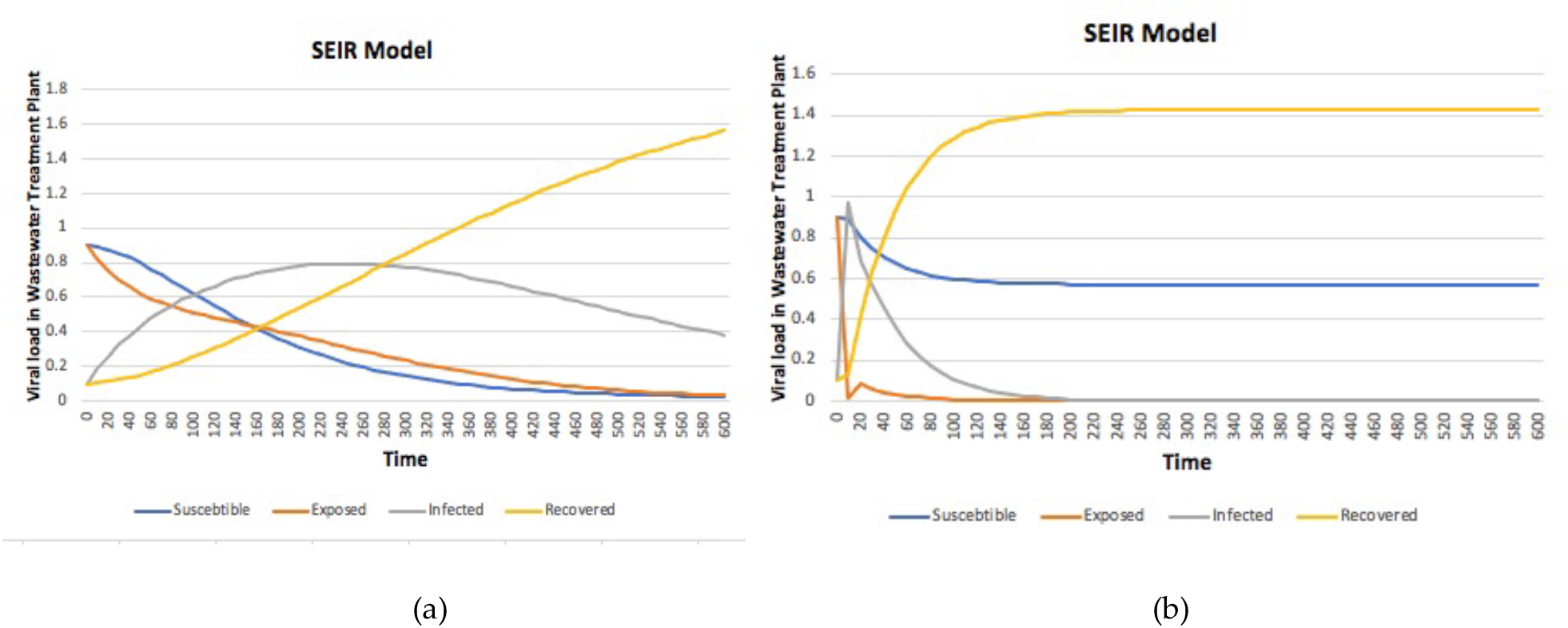
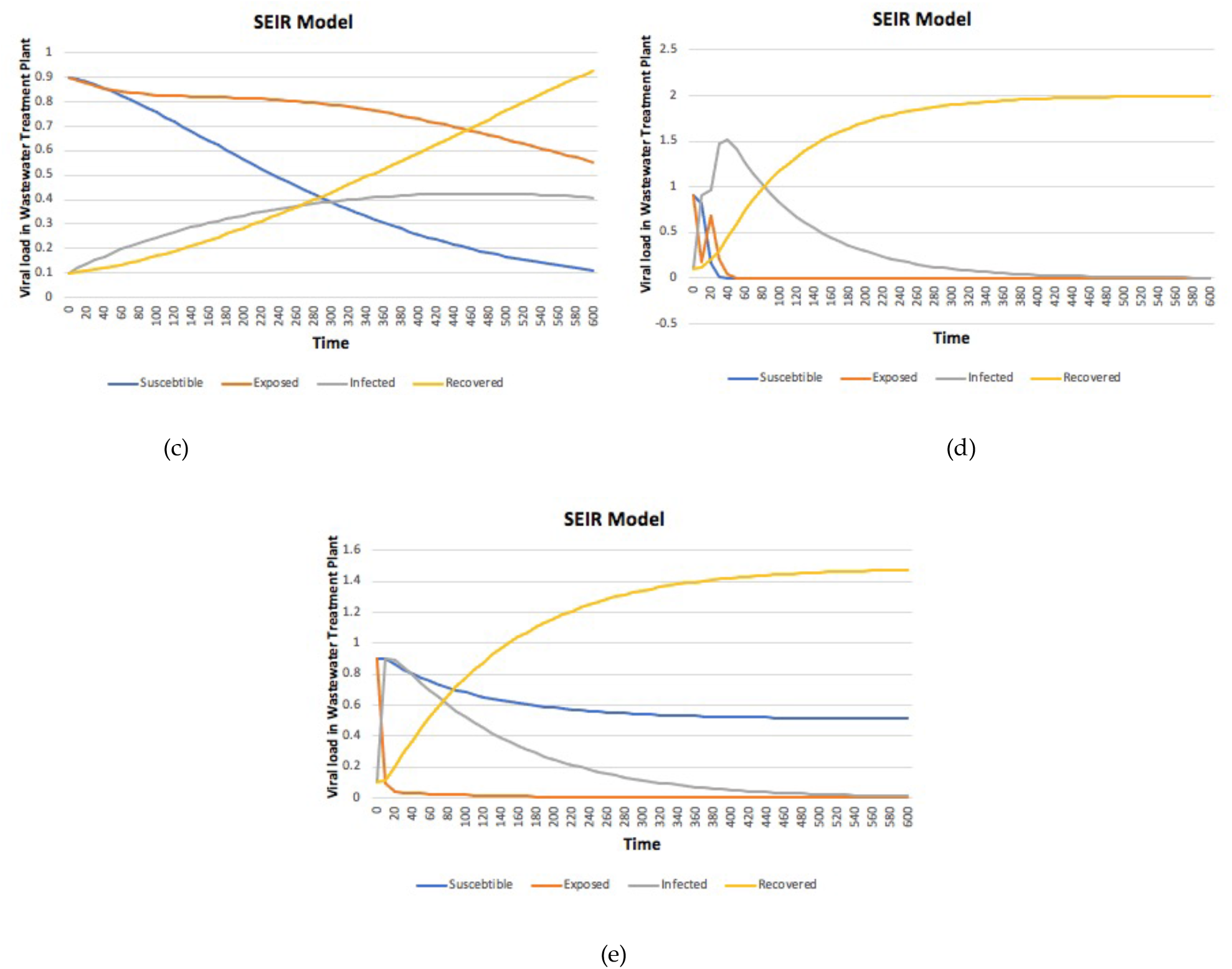
Numerical solution for SEIR mode with (a) ***β*** = **0. 0095, *γ*** = **0. 004, *σ*** = **0. 01**, (b) ***β*** = **0. 0095, *γ*** = **0. 03, *σ*** = **0. 1**, (c) ***β*** = **0. 01, *γ*** = **0. 004, *σ*** = **0. 0025**, (d) ***β*** = **0. 09, *γ*** = **0. 01, *σ*** = **0. 09**, (e) ***β*** = **0. 00348, *γ*** = **0. 03, *σ*** = **0. 1**.

### 4.3 Gaussian Model

The Gaussian model was used to predict the time evolution of the COVID-19 pandemic in Germany [25], Italy [26], and China [27]. The best justification for the Gaussian distribution for detecting the dynamic behavior of SARS-CoV-2 is given by the central limit theorem. For instance, when several independent random variables are added, their normalized sum satisfies the Gaussian distribution, even if the original variables are not normally distributed. Numerical simulations of earlier pandemics indicate that the time evolution of pandemic waves is usually shown by an exponential rise until a maximum is reached and then decreases rapidly. We adopted a simple Gaussian distribution to estimate the dynamic nature of coronavirus infection in wastewater. Let *X*_*i*_(*t*) be the number of viral loads in an untreated wastewater plant per day, and its time evolution is given by the Gaussian function:

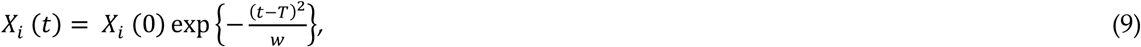

shown in Figure 5(b), where *X*_*i*_ (0) denotes the maximum value at a certain time *T*, and *w* is the width of the Gaussian. By monitoring the new viral loads in wastewater and using equation (6), we can derive the relative change in COVID-19 infection per day as follows:

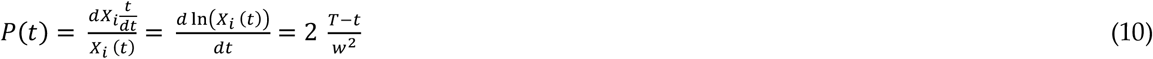

where ***P***(***t***) denotes the Gaussian distribution of the viral load of coronavirus traces in untreated wastewater. The monitored data are often reported in terms of the doubling time (***τ***)and the effective reproduction factors. The corresponding exponential function at any time is given as:

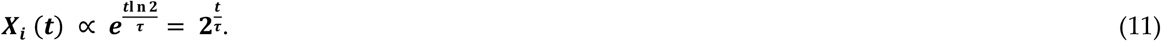

Using Equation (8) in Equation (7), we obtain the relative changes in the daily rate:

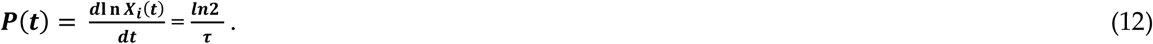

Equating the two results in Equations (7) and (9), we obtain the time-dependent Gaussian doubling time:

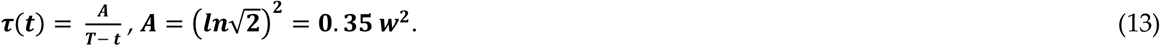

The daily number of newly infected cases can also be defined using the cumulative case rate. Using Equation (10) and the peak time ***t*** − ***T***, the corresponding cumulative number of cases at time ***t*** in terms of the error function can be written as

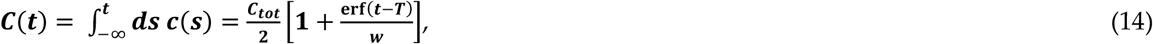

where 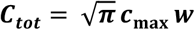 denotes the total number of infected cases. Similar values for cumulative death and infection cases relevant to the first wave of the COVID-19 pandemic were obtained in [28]. Using Equation (14), we can write the respective relative changes as follows:

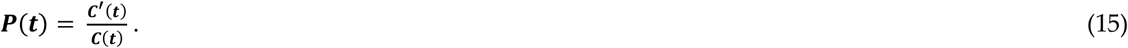

Using Equation (9) and equating the results with the corresponding exponential function leads to the time-dependent cumulative Gaussian doubling time:

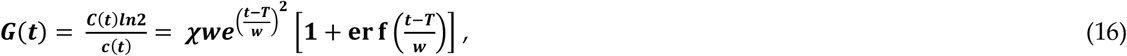

where 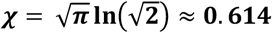. Figure 5(a) shows the Gaussian instantaneous time and cumulative doubling time as a function of time.

The Gaussian model can predict the time (months) evolution of the number of infections, where ***X***_***i***_ (**0**) denotes the maximum number of infections. The best-fit parameters have a peak time ***T*** = **4. 4** months, where ***t*** = **0** in ***X***_***i***_ (***t***) corresponds to March 2, 2020 (the first day of COVID-19 infection in Saudi Arabia). The maximum infection cases were observed during June and July 2020, approximately 4.4 months after the first day of infection in Saudi Arabia.

### 4.4 Susceptible Exposed Infectious Recovered (SEIR) model

The simulation of the well-known SEIR model was performed to represent the dynamic behavior of the spreading process of the COVID-19 pandemic through wastewater. In epidemiology, the SEIR model is a compartmental pandemic model widely used to characterize the outbreak of COVID-19. In this model, the spread of infection depends on the number of susceptible populations and the number of infected populations. The incubation period is considered when individuals have been infected but do not show symptoms. Because the coronavirus disease has a long incubation period, it is reasonable to model the pandemic with another compartment that is exposed to humans, but not virus spreaders [29].

In the SEIR model, we assume that the time is long term with no vital dynamics, and the population *N* size is constant. In this model, individuals are classified into four groups or compartments according to their infectious status. We classified the infection types as either with or without symptoms. The compartments of the model are as follows:

- Susceptible (*S*): number of humans that are not infected by the virus but may catch the disease.
- Exposed (*E*): number of humans that are already infected but cannot spread the virus.
- Infected (*I*): number of infected humans and transmission of the virus to susceptible individuals.
- Recovered (*R*): number of humans that are recovered from infection.

A simple model of SEIR is given as:

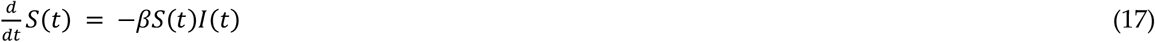

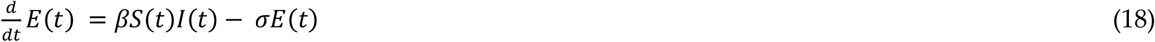

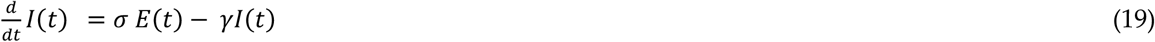

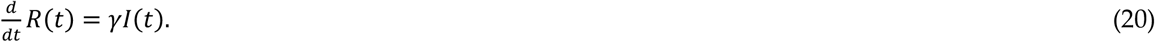

The above system of equations (17–20) can be solved using any numerical method, and a simulation can be presented to predict the behavior of SARS-CoV-2. The above equation can be written as follows:

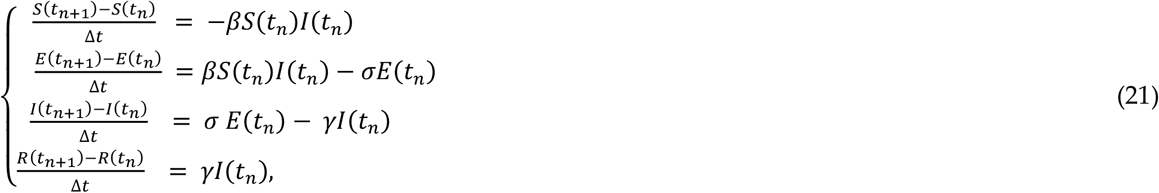

where Δ***t*** represents a very small step size in the time domain. The parameters ***β, σ***, and ***γ*** defined in the above equations represent the probability of infection, incubation rate, and average rate of people recovered at time ***t*** (in days), respectively. We assumed the average incubation period to be five days.

The number of possible viral loads in the wastewater plants can be predicted, as shown in Figure 6(a)–(e). From the model diagram, we can see that the population of infectious individuals increases at the beginning of the outbreak and progressively decreases over time. The main feature of this model is the incorporation of the importance of coronavirus contamination in untreated wastewater by infected people.

## 5. Results and Discussion

The possible COVID-19 hotspots in different wastewater treatment plants of five major cities in Saudi Arabia can be predicted using log-normal mathematical models and a Gaussian model. The results are shown in Figures 4(a)–(b) and 5(a)–(b), respectively. Moreover, further prediction of the next hotspot was considered using the susceptible-exposed-infected-recovery (SEIR) mathematical model. The results for predicting possible viral loads in wastewater treatment plants are shown in Figure 6(a)–(e).

We can assume that the viral load threshold is when the viral load in a wastewater treatment plant is equal to 1. From Figure 4(a), it can be concluded that the regions that are at maximum risk from COVID-19 infection are from untreated wastewater plants with sharply peaked curves. The black color curve shown in Figure 4(a) indicates a high number of COVID-19 infection cases on the first day, while the last region with a red color curve has a continuous number of moderately infected persons. However, as shown in Figure 4(b), the data collected from the wastewater plants were assumed to follow a log-normal distribution. It is presumed that data were collected from wastewater treatment plants from five major cities in Saudi Arabia with daily averages and standard deviations in the case of the Gaussian model. It can be observed that wastewater treatment plants with black and green curves are considered as COVID-19 hotspot regions. The remaining curves were below the viral load threshold. In particular, we can conclude that these two regions, Makkah (average = 0.2, standard deviation = 0.1) and Riyadh (average = 0.3, standard deviation = 0.4) have a higher number of infected persons with SARS-CoV-2. The prediction of hotspots using a log-normal model is consistent with the real data of COVID-19 infection cases observed in the major provinces of Saudi Arabia. In March 2020, the province of Makkah had the highest number of COVID-19 infection cases (30.7%), followed by Riyadh (23%) [55]. In general, we can compare all the collected data from the wastewater treatment plants weekly or monthly, and each region can define a threshold. A COVID-19 hotspot region can be identified from a wastewater treatment plant with a viral load above the threshold value.

The Gaussian model can also be used to predict the possible peak time of infection using data collected from wastewater treatment plants. From Figure 5(b), the maximum probability of infection cases occurs during the time ***T*** = 4.4 (months), which is approximately four and a half months since the first day of the COVID-19 infection case in Saudi Arabia. The cumulative Gaussian model predicts the possible viral load in wastewater treatment plants over time. According to the model, the cumulative cases of infected people are represented by the red curve. This signifies that with a moderate rate of viral load in a wastewater treatment plant, there is a possibility of the occurrence of COVID-19 over a long period.

Using the epidemiological mathematical model SEIR and simulations can predict the next COVID-19 hotspot region. The model also shows a decrease in the susceptible population as people become exposed and infected and then recover. The coronavirus load per day in untreated wastewater was predicted for five different regions, and Figures 6(a)–(e) show the numerical simulations of the model with infection, incubation, and recovery rates for different regions in Saudi Arabia. From the simulation process, the SEIR model with infection type had a higher prediction of the hotspot. Figure 6(d) represents the hotspot because we assume that the threshold value for the SARS-CoV-2 virus load in an untreated wastewater treatment plant is 1. We can see that the viral load in untreated wastewater is higher and declines progressively over time.

All mathematical model approaches mentioned above may be promising methodologies for predicting possible COVID-19 hotspots. However, our results indicate that traveling restriction, social distancing, vaccination, medical intervention, and security are important measures to flatten the curve.

## 6. Conclusions

In this study, we sought to identify COVID-19 hotspots in wastewater collected from wastewater treatment plants in major cities in Saudi Arabia. SARS-CoV-2 is spread not only through the air but also through untreated wastewater, reaching wastewater treatment plants and the aquatic environment. Therefore, significant attention is required to examine wastewater treatment plants, especially in Saudi Arabia, where only a part of the wastewater is treated. We highlighted recently developed wastewater treatment plants based on virus models that can be used to differentiate virus removal efficiencies. This includes some advanced techniques, such as treatment using ozone, UV, ultrafiltration, and nanofiltration membrane-based systems. It is a combination of different techniques in primary, secondary, and tertiary treatments that would facilitate the viral removal treatment process in wastewater treatment plants.

We used mathematical modeling, including the Gaussian model, the log-normal model, and the SEIR model to understand the possible coronavirus load in untreated wastewater from wastewater treatment plants. The COVID-19 hotspot was predicted using these models, where the collected data were plotted as a function of time. The hotspots were identified when the viral load of the SARS-CoV-2 trace in wastewater was higher than the threshold value. Using the log-normal model to predict the region with the maximum infection of COVID-19 through wastewater provides a promising mathematical model. This model approach predicts two provinces (Riyadh and Makkah) in Saudi Arabia as COVID-19 hotspots, which is close to the real data of infection. The Gaussian model aids to estimate the time of the possible maximum outbreak of the pandemic. It was predicted that the peak outbreak would be 4.4 months since the first day of the outbreak in Saudi Arabia, which is consistent with the real data because June and July 2020 recorded the maximum number of infections. To describe the possible viral load in a wastewater treatment plant using the SEIR model has more advantages in the long-term prediction of the nature of the COVID-19 virus. In conclusion, our models follow the process of predicting COVID-19 hotspots in wastewater. Wastewater treatment plants could be one of the solutions to limit the viral spread into the environment and a possible future detection, along with accessing the Saudi Health Ministry control measures and decisions on infection.

## Data Availability

All data produced are available online at the website of Ministry of Health Saudi Arabia.

https://covid19.moh.gov.sa/

## Acknowledgments

The authors would like to thank Scientific Research Ethics Committee, Jazan University, Ministry of Higher Education, Saudi Arabia, for financial support of this research with grant number W4-059.

